# Temporal and spatial analysis of *Plasmodium falciparum* genomics reveals patterns of connectivity in a low-transmission district in Southern Province, Zambia

**DOI:** 10.1101/2021.10.14.21264576

**Authors:** Kara A. Moser, Ozkan Aydemir, Chris Hennelly, Tamaki Kobayashi, Timothy Shields, Harry Hamapumbu, Michael Musonda, Ben Katowa, Japhet Matoba, Jennifer C. Stevenson, Douglas E. Norris, Philip E. Thuma, Amy Wesolowski, William J. Moss, Jeffrey A. Bailey, Jonathan J. Juliano, on behalf of the Southern and Central African International Center for Excellence in Malaria Research (ICEMR)

## Abstract

Understanding temporal and spatial dynamics of ongoing malaria transmission will be critical to inform effective interventions and elimination strategies in low transmission regions approaching elimination. Parasite genomics are being used as a tool to monitor epidemiologic trends, including assessing residual transmission across seasons or importation of malaria into these regions. Southern Province, Zambia is a low-transmission setting with seasonal malaria. We genotyped 441 *Plasmodium falciparum* samples using molecular inversion probes at 1,832 positions across the genome, using dried blood spots collected from 2012-2018 from 8 health centers in the catchment area of Macha Hospital in Choma District. We show that highly related parasites persist across multiple seasons, suggesting that the persistence of malaria is at least in part fueled by parasites “seeding” across the dry season. In addition, we identify clusters of clonal parasites that are dissimilar to the general population, suggesting that introduction of parasites from elsewhere may contribute to the continued malaria burden. We identified signals of population size fluctuation over the course of individual transmission seasons, suggesting a ramp-up of malaria transmission from a season’s beginning. Despite the small spatial scale of the study (2,000 sq km), we identified an inverse relationship between genetic relatedness of parasite pairs and distance between health centers, as well as increased relatedness between specific health centers. These results, leveraging both genomic and epidemiological data, provide a comprehensive picture of fluctuations in parasite populations in this pre-elimination setting of southern Zambia.

## INTRODUCTION

As malaria elimination efforts continue to drive down disease burden in Africa, regions previously endemic for malaria have seen drastic reductions in overall morbidity and mortality (Björkman et al., 2019; Masaninga et al., 2013). In these areas now nearing elimination, identification of ongoing local transmission and importation events is critical for maintaining elimination gains and preventing outbreaks in increasingly immune susceptible populations. Identifying reservoirs for continued transmission, which may occur in specific geographical locations, temporal periods, or age groups would allow targeted control efforts to interrupt sustained transmission. Identifying suspected imported cases (where a genetically distinct parasite is introduced to a region) would assist malaria control programs in identifying high-trafficked routes of human movement and sources of importation (Tessema et al., 2019). Blocking such cases would also be key to protect against introductions to populations which are increasingly more susceptible to malaria after successful control efforts (Mundagowa and Chimberengwa, 2020; Patel et al., 2014).

Genetic epidemiology can be used to track ongoing transmission patterns in pre-elimination settings and help elucidate the mechanisms of persistent transmission (**Figure 1**). Following decreases in pathogen transmission, it is expected that parasite population sizes will also shrink, reflected in reduced genetic variation fueled by higher levels of clonal transmission and inbreeding (Branch et al., 2011; Daniels et al., 2015; Nkhoma et al., 2013). These genetic patterns should follow reductions in disease burden over the long-term, but may also be seen in areas with distinct seasonality with low transmission periods (e.g. dry seasons). These expectations of low-genetic variation in association with low transmission may not hold if an area receives high levels of imported cases (Raman et al., 2020; Roh et al., 2019). Additionally, these regions may be more sensitive to temporal fluctuations in terms of malaria cases and parasite populations, as increased transmission and outbreaks might be more common due to decreasing immunity (Obaldia et al., 2015).

**Figure 1:**
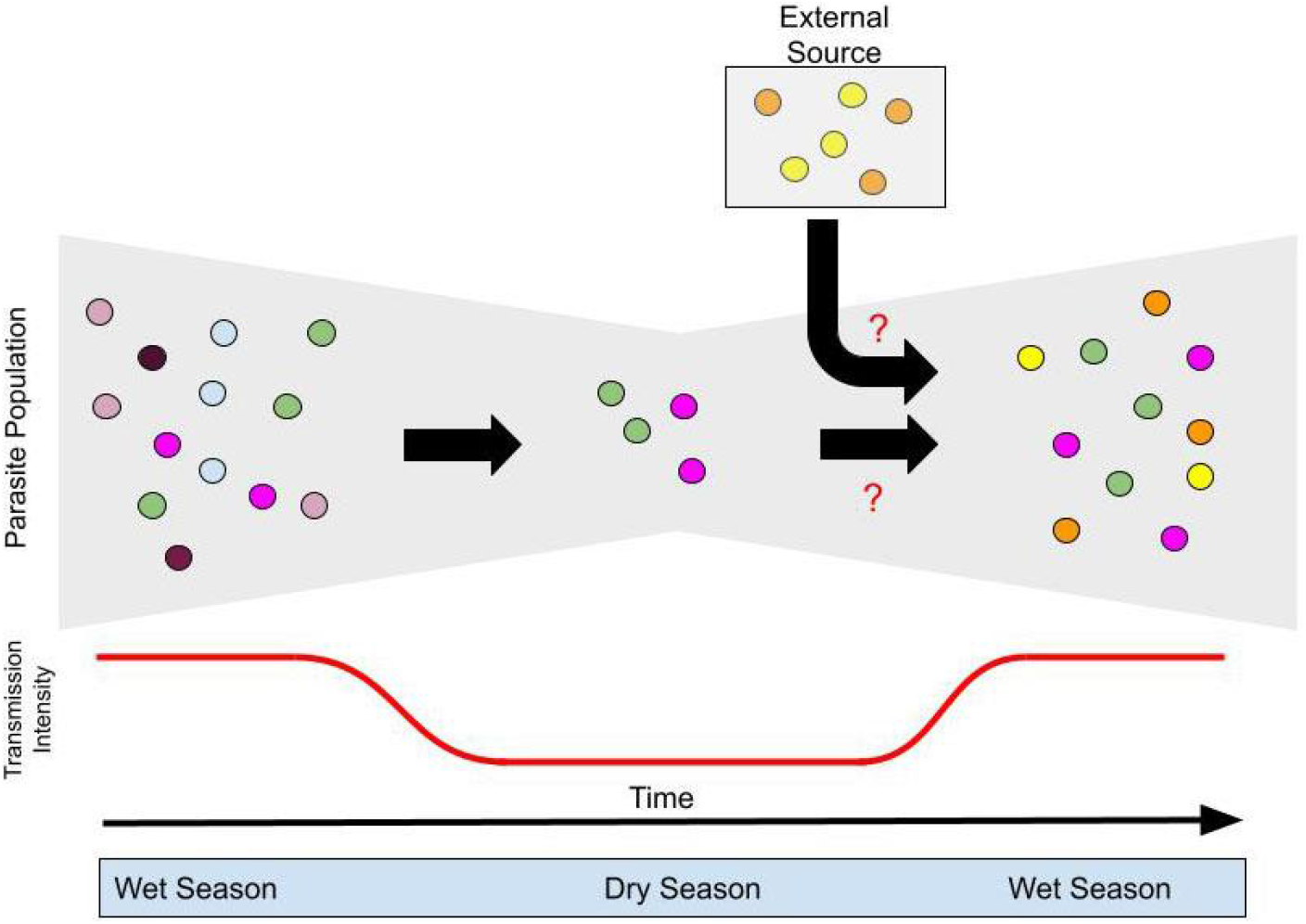
Theoretical construct for continued transmission in low transmission settings. Malaria transmission in a low endemicity setting is likely the result of a combination of persistence through the dry, low transmission season and importation from other regions. However, the relative contribution of these remains unknown (red question marks). Parasite genomics can help understand these relationships. Different parasite lineages (colors of circles) may or may not survive through a dry season, shown by the reduction in diversity during the dry season. However, genetic diversity may be enhanced through importation through an external source.

These low transmission settings offer a unique opportunity to observe long-term dynamics of parasites across seasons, as the smaller number of parasites may be easier to track from a lack of recombination with genetically distinct parasites within the mosquito. A small number of studies have attempted to observe temporal population changes in these regions, and even fewer have been able to combine temporal and geospatial data to identify hotspots over space and time (Chenet et al., 2012; Daniels et al., 2015; Gwarinda et al., 2021; Kattenberg et al., 2020; Noviyanti et al., 2015; Roh et al., 2019; Tessema et al., 2019). Most of these studies evaluated parasite population changes over relatively large geographic distances. Thus, it remains relatively unknown if and how parasite populations change over time in a low transmission setting on a small regional scale, such as a district.

Southern Zambia has seen a drastic reduction in the number of malaria cases over the past two decades (Masaninga et al., 2013). Compared to the turn of the century, morbidity and mortality in this region is low (Ministry of Health, n.d.). Nevertheless, Choma District consistently continues to have a low number of malaria cases every year, and these tend to occur seasonally (Kobayashi et al., 2019). However, it is not understood how the underlying parasites are maintained between seasons. Here we use a novel high throughput genomics tool, molecular inversion probes (MIPs), to characterize the transmission dynamics that connect falciparum malaria infections across space and time in Choma District, Southern Province, Zambia from 2012-2018. This approach allows us to develop a more refined understanding of transmission and importation dynamics than previous studies using low resolution genotyping as we can use identity-by-descent (IBD) approaches to evaluate relationships between parasites. IBD has been shown to be superior for understanding interconnectivity of parasite populations (Henden et al., 2018; Schaffner et al., 2018; Taylor et al., 2017). We show that highly related parasites are connected across multiple seasons, suggesting that cases are at least in part fueled by clonal lineages persisting through the dry season. In addition, we identify clusters of clonal parasites that are dissimilar to the general population, suggesting that introduction of parasites from elsewhere may contribute to the continued malaria burden.

## RESULTS

The epidemiology of malaria in the Matcha Hospital catchment area within the Choma District from 2012-2018 was seasonal (**Figure 2**). Malaria cases started to increase around December of each calendar year, roughly peaking in May the following year. Little-to-no cases were reported from July through November. 441 DBS samples were collected at eight rural health centers and Macha Hospital (**Figure 2**) during this time period and 302 (68%) were successfully genotyped at 1,410 positions across the genome. These samples are representative of almost all time periods from which the original sample set were collected (**Figure S1**). 84% of samples (n=255) had less than 10% missing genotype calls across all positions (**Figure S2**). Most successfully genotyped samples were collected from 2016 and 2017, but samples representing all years between 2013 and 2018 were also successfully genotyped and reflect the epidemiologic case data (**Figure 2**).

**Figure 2:**
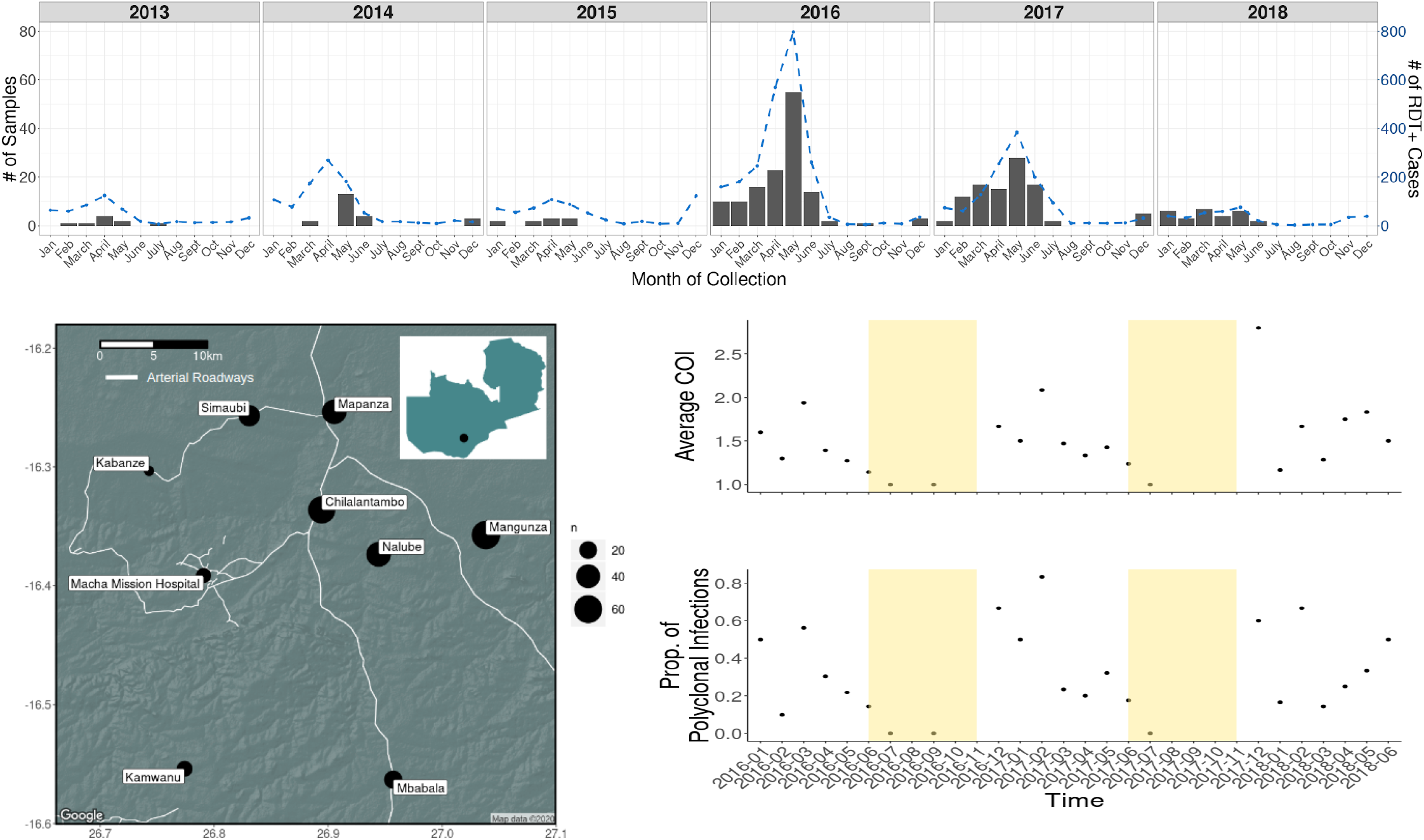
Malaria trends and location of sample collection. The top panel shows trends of RDT positive cases at the 8 health centers over the course of the study (blue dotted line) and the number of dried blood spot (DBS) samples collected by month over the course of the study (grey bars). The lower left panel shows the locations of the health centers in the district with the size of the mark representative of the number of samples collected at that clinic. The bottom right panel shows the average complexity of infection (COI) and proportion of polyclonal samples genotyped each month with the dry season marked by the yellow bar.

Genetic data generated through MIPs showed signs of both increased transmission and parasite population growth across seasons, expected in regions that have seasonal trends. Complexity of infection (COI) often tracks with transmission intensity on populations (Hendry et al., 2021; Verity et al., 2019; Watson et al., 2021). Estimates of COI are shown in **Figure 2**. The majority (67%, n=202) of infections were monoclonal, as expected from an area with overall low transmission (distribution of all COI estimates can be found in **Figure S3)**. However, average COI estimates increased with periods of time coinciding with the rainy season (roughly November to May of each year), with the lowest average COI estimates overlapping with the dry season.

Genetic relatedness was calculated between all infection-pairs using the inbreeding coefficient *F* [as described in (Verity et al., 2019)]. Despite the overall low-transmission setting, the majority of comparisons showed little-to-no genetic relatedness (**Figure 3**). However, 6% of comparisons had an inbreeding coefficient > 0.25. Infection comparisons collected from the same health center were more likely to have higher *F* values. Additionally, a rough isolation-by-distance trend was observed between comparisons across different health centers (**Figure 3**). To identify if hotspots of transmission of highly-related infection pairs were occurring between certain health centers, the number of highly-related (*F* > 0.25) infection pairs occurring between two health centers was shown as a proportion of the total number of highly related pairs (**Figure 3**). Visual inspection of these trends showed that health centers connected by major roads were more likely to have more closely related parasites to one another than those not connected by major roadways (such as Kamwanzu), although geographic proximity could override this pattern (ie, Nalube). However, all health centers, even those geographically distant or not connected by major roadways, contained highly-related infection pairs.

**Figure 3:**
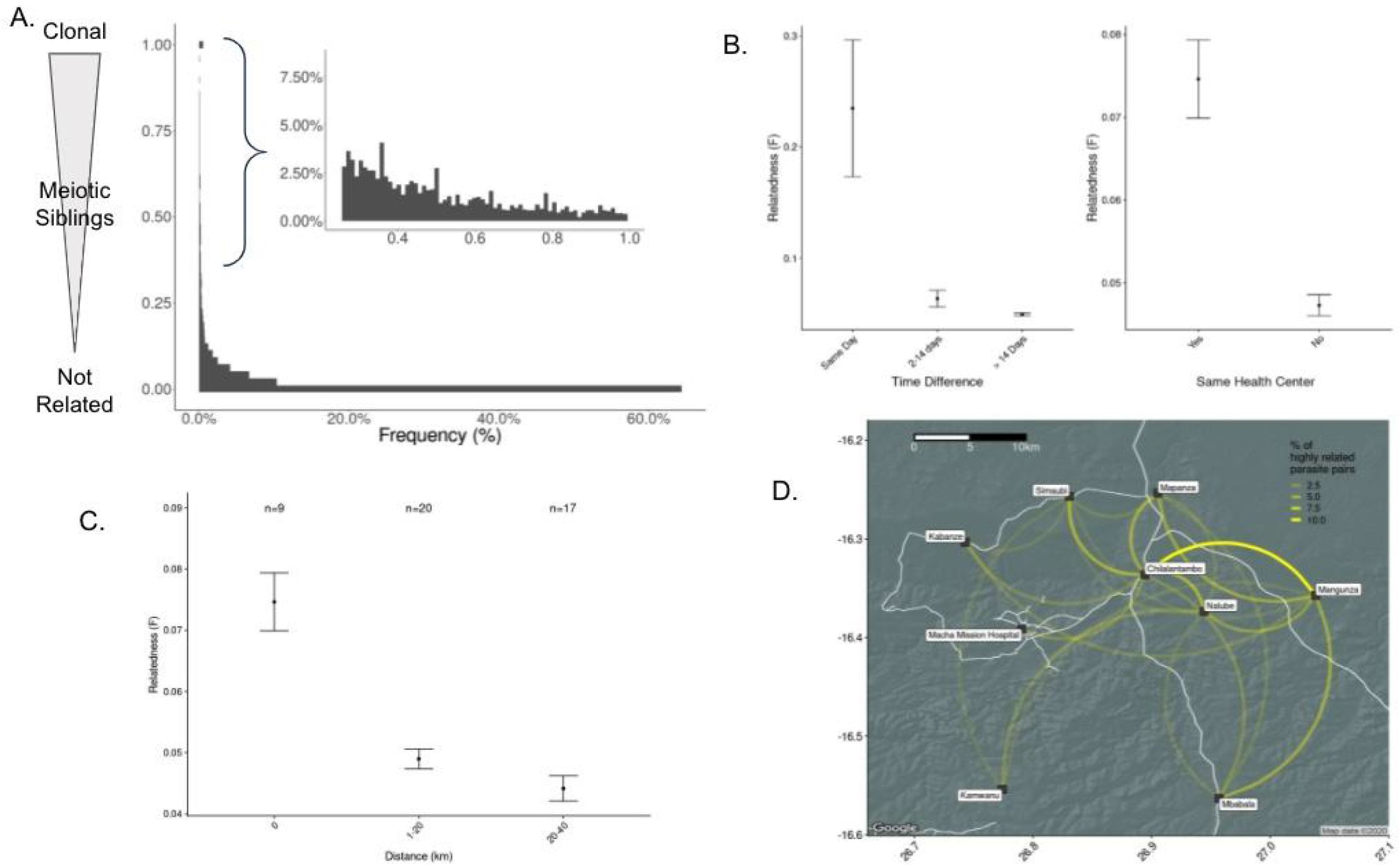
Temporal and spatial patterns in pairwise genetic relatedness. Panel A shows the genetic relatedness between all samples using the *F* statistic. The insert shows the number of samples with the highest relatedness. Panel B shows that isolates collected on the same day and within the same health center were more likely to be highly related. In addition, Panel C shows a pattern of isolation by increasing geographic distance. Panel D shows the proportion of highly related parasite pairs shared between health centers. The width of the yellow connecting line corresponds to the proportion of shared highly related parasites. Major roads connecting health centers are shown in white.

Similar to the observed spatial trends above, infections collected on the same day had higher *F* values than samples collected further apart in time (**Figure 3**). To further investigate temporal patterns of genetic relatedness, networks were built using monoclonal infections. Interestingly, these networks showed that there were highly-related (*F* > 0.25) infection pairs that occurred across months of the same malaria season, and even between malaria seasons (**Figure S4**). Using a cut off of F>0.25, the equivalent of half-siblings or closer relationships, we identified networks of monoclonal infections involving 38 isolates across multiple seasons that also contained infections sampled during the dry season (n=5) (**Figure 4**). This provides evidence that a proportion of the parasite population is maintained through the dry season and contributes to cases of disease during the following season. The largest cluster traverses multiple dry seasons, indicative of long-term propagation with minimal outcrossing in a proportion of parasites. Among clonal samples (*F* = 1, n=104), multi-season networks or those spanning multiple months occurred (**Figure 5**), including three clusters comprising more than seven samples each. These clusters all represent samples collected from the 2015-2016 and 2016-2017 seasons, the seasons for which most samples were successfully analyzed. Interestingly, these clonal networks showed little spatial clustering with clones appearing across different health centers (**Figure 5**).

**Figure 4:**
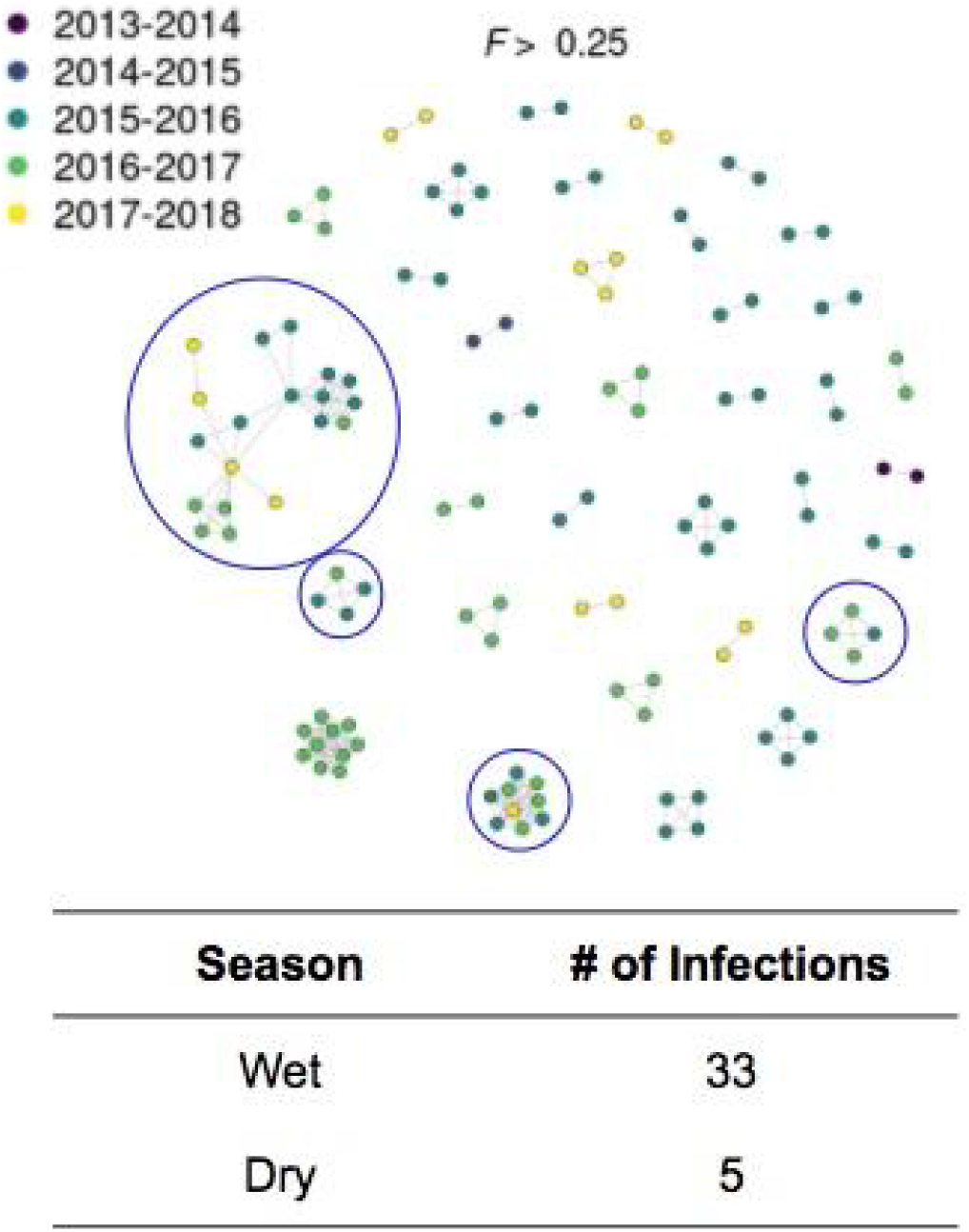
IBD network of monoclonal samples with *F* > 0.25 shows transmission across multiple seasons and through the dry season. Networks spanning multiple seasons are circled in blue. The samples are colored by the season of collection. Thirty-eight samples were contained in these networks, with five representing samples collected in the low transmission dry season.

**Figure 5:**
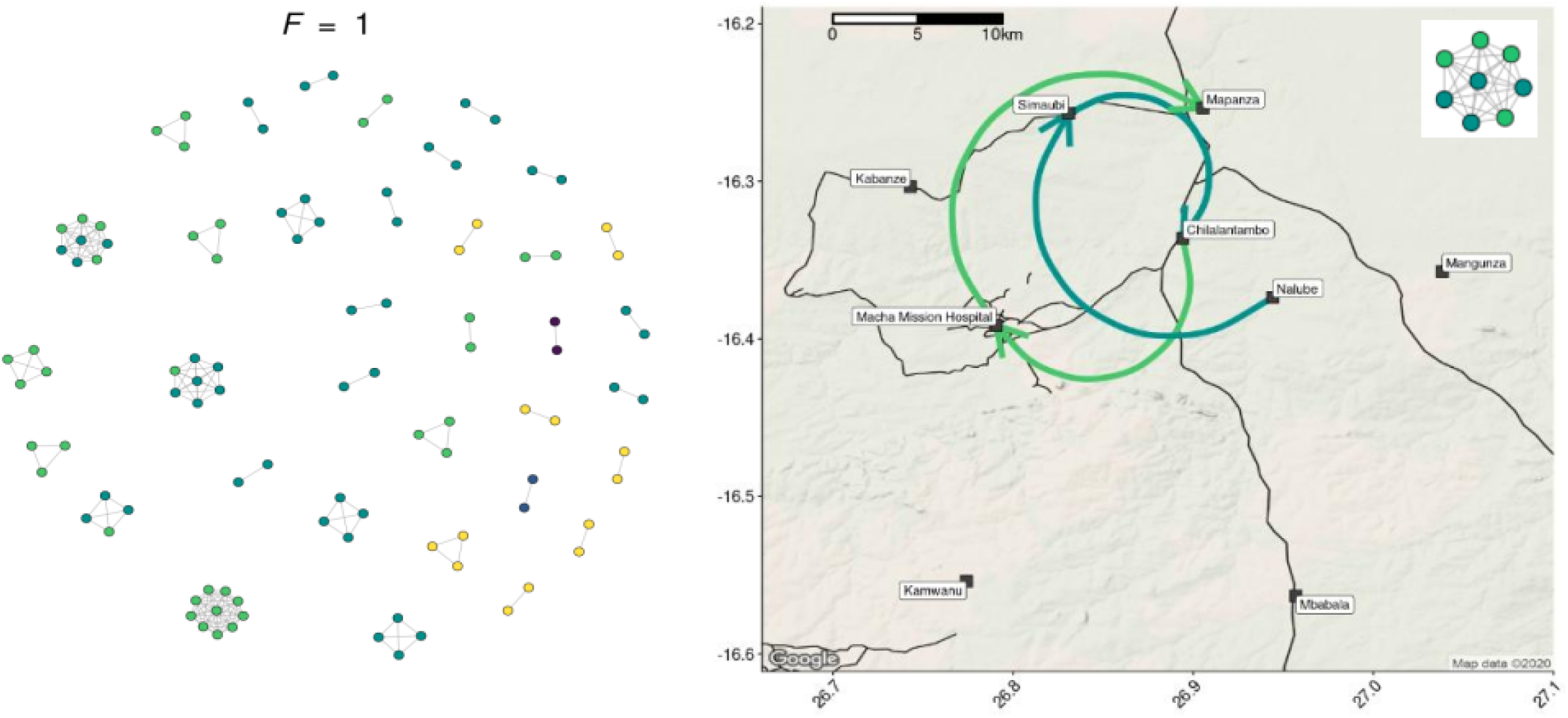
IBD networks of clonal samples (*F* =1) show transmission across seasons and across the study site. In the left panel, networks across seasons are documented using only monoclonal samples. The right panel shows the geographic locations of samples from one of these clusters. These locations are connected by lines colored to correspond to the season of collection. Arrows on these lines correspond to the order in which the samples were collected. The insert represents the cluster from the left panel displayed on the map.

While these findings provide evidence that parasites are maintained through the dry season to cause disease in the subsequent malaria season, it does not address the potential contribution of imported parasites. To investigate if genetic outliers existed among the infections presented here (a possible proxy for importation of genetically distinct parasites), a principal component analysis (PCA) was conducted using all infections (**Figure 6**). Overall the first two components explained very little of the variation in the dataset (PC1: 4%, PC2: 3%). However, PC1 and PC2 separated a small number of infections from all other infections. Interestingly, these infections were two of the large *F*=1 clusters reported above.

**Figure 6:**
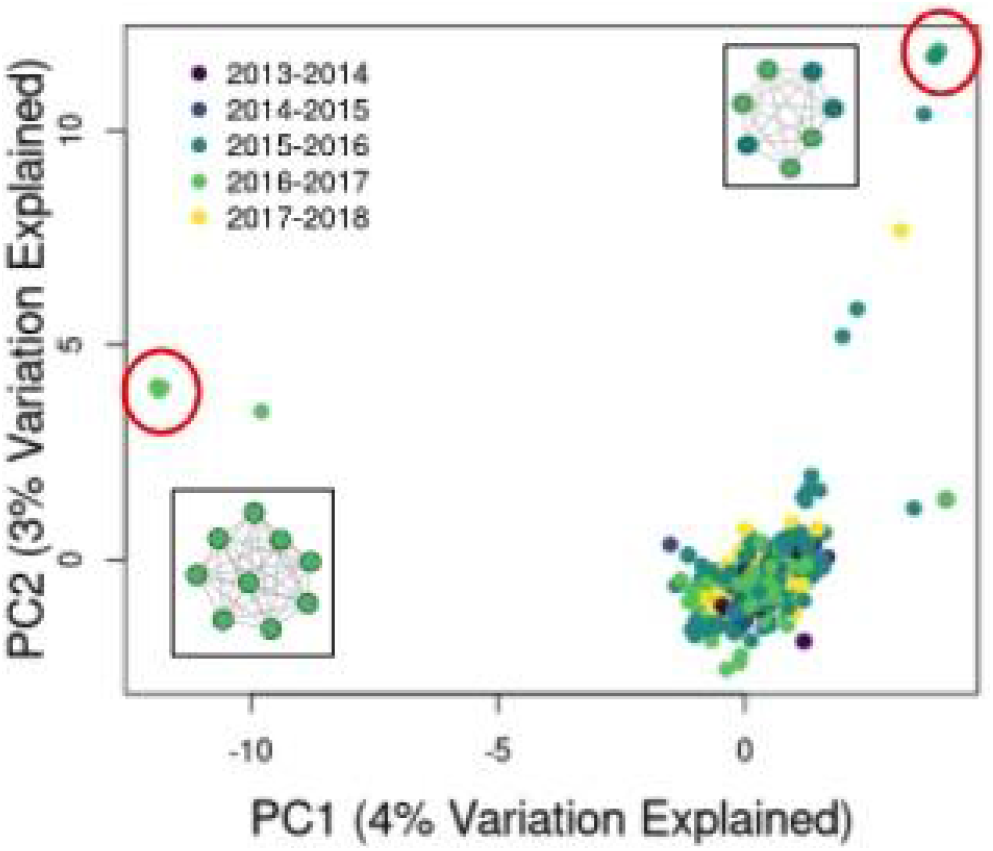
Principal coordinate analysis (PCA) of parasite isolates suggests clonal expansions of the least related parasite genotypes. The PCA components 1 and 2 of all isolates show several parasite samples that appear genetically different than most samples in the study. Two clusters (circled in red) represent two of the networks of clonal isolates (inserts) previously identified (**Figure 5**).

## DISCUSSION

Using a novel high-density genome-wide genotyping platform, we were able to provide a detailed snapshot of the transmission dynamics that connect falciparum malaria infections across space and time in the Choma District of Zambia from 2012-2018. Despite the relatively small geographic region in the study (2,000 sq km) we were able to determine that the majority of infections were monoclonal and networks of identical or highly related parasite persisted across space and time and were detected in the dry low transmission season followed by “seeding” the parasite population in the next high transmission season (**Figure 4** and **5**). These parasites were then transmitted widely across the district, likely due to the relatively small geographic scale. These transmission networks appear to overlay a background population that maintains an overall weak signal of isolation by distance (**Figure 3**), suggesting that these networks were potentially acting like small outbreaks on top of the general parasite population. Interestingly, two of the clonal networks were among the most dissimilar parasites to the general parasite population, suggesting that their origin may have been outside of the general study area (**Figure 6**). This suggests that importation with resultant local spread may have led to these clusters and supports the theoretical construct for persistence of parasite populations across the dry season in Choma District (**Figure 1**). To fully define the role of importation we would require genetic information from contemporary parasite populations from outside Choma District, as well as information on human migration.

Genomic studies of local transmission and importation in low and medium endemicity countries are becoming more common in the literature and starting to shed light on the dynamics of parasite movement (Chenet et al., 2012; Daniels et al., 2015; Gwarinda et al., 2021; Kattenberg et al., 2020; Morgan AP, Brazeau NF, Ngasala B, Mhamilawa LE, Denton M, Msellem M, Morris U, Filer DL, Aydemir O, Bailey JA, Parr JB, Mårtensson A, Bjorkman A, Juliano JJ, n.d.; Noviyanti et al., 2015; Roh et al., 2019; Tessema et al., 2019). However, relatively few studies have used genomic approaches to study malaria transmission dynamics and importation on small geographic scales, such as a single district (Searle et al., 2020). Detailed work in Senegal has shown similar findings, including persistence of clones across multiple seasons and evidence of importation into similar low transmission regions (Daniels et al., 2020, 2015; Sy et al., 2020). However, the approaches used in these studies have been low density genotyping, primarily a 24 single nucleotide polymorphism (SNP) barcode. This study adds to the existing literature as the high density genotyping allows for the use of IBD to help resolve finer structure and relationships within the population.

Unlike previous work (Searle et al., 2020), we were able to use IBD to evaluate networks of parasites based on the number of outcrossing, showing that networks of half-siblings or more highly related parasites are extensive and persist for longer than networks of clonal parasites (**Figures 4** and **5**). In addition, we were able to use this metric to define the underlying isolation-by-distance relationship seen in the general parasite population in Choma District, allowing us to hypothesize about the relative contributions of imported and endemic parasites. In addition, as noted previously, the MIP approach provides a cost-effective means of high resolution genotyping of large numbers of parasites compared to whole genome sequencing and is therefore a scalable tool to provide detailed studies of transmission and importation with large numbers of samples (Aydemir et al., 2018). Lastly, within the study region, we were able to detect fluctuations in transmission intensity, as measured by COI, between seasons, suggesting a ramp up and expansion of parasite populations with each season. COI has previously been shown to be correlated with local transmission intensity (Hendry et al., 2021; Verity et al., 2019; Watson et al., 2021).

While this work demonstrates maintenance of parasite clones through the dry season and supports the high likelihood of importation of clones into the district that fuel small outbreaks, there are multiple limitations. First, the sampling of only acute cases will limit the ability to define local transmission dynamics and identify importations. The asymptomatic reservoir likely contributes significantly to sustained low level transmission (Lin et al., 2014). However, given the low density parasitemias of asymptomatic infections, genotyping of these parasites remains a challenge with any platform. Second, we did not obtain detailed travel histories on the participants, making it impossible to determine if the genetically unrelated parasites are imported. Lastly, the relatively small sample size prevents us from quantifying the relative burden of importation versus persistent transmission over the dry season on the parasites that dominate in the following high transmission season. This will require further studies, requiring dense sampling of symptomatic and asymptomatic cases with travel histories.

The success of malaria elimination in low transmission regions in Africa will depend on our understanding of transmission dynamics and importation. Per the World Health Organization, defining imported vs. locally acquired cases is critical for designing and stratifying targeted interventions (Marshall et al., 2016; World Health Organization, n.d.). This work demonstrates the feasibility of the use of genome-wide approaches to help define the relationships between infections in settings approaching malaria elimination. The genotyping tools and analytical methods for these studies are continually advancing. Although we focused largely on monoclonal samples, significant information likely can be gathered from addressing polygenomic infections. Preliminary work suggests that these tools will still be effective in studying parasite populations in high transmission settings and can reveal important patterns of transmission (Verity et al., 2019). In addition, combining human mobility data with these genomic tools will help us better understand both local transmission and importation. This study leveraged state of the art genomic tools and analysis methods to provide a detailed snapshot of the transmission dynamics of malaria in a region on the cusp of elimination and highlights the feasibility of these methods to inform targeted interventions to achieve and sustain malaria elimination.

## MATERIALS AND METHODS

Samples were collected through passive case detection from eight health centers in and around the catchment area of Macha Hospital in Choma District, Southern Province, in an area of 2,000 km^2^. Total number of cases with positive RDTs at these clinics were tracked during this period. This work was approved as part of ICEMR study by the Tropical Diseases Research Centre, Ndola, Zambia (Ref No: TDRC/ERC/2010/14/11) and Johns Hopkins Bloomberg School of Public Health (IRB # 3467). Analyses utilizing parasite genomes from de-identified samples were deemed nonhuman subjects research at the University of North Carolina at Chapel Hill (NC, USA) and Brown University (RI, USA).

From 2012-2018, dried blood spots (DBS) were collected from 441 individuals presenting to the health centers with symptoms who were positive for *P. falciparum* infections by rapid diagnostic test. DNA was extracted from each DBS with a Chelex-Tween protocol (Topazian et al., 2020). Parasitemia was assessed using quantitative PCR with probes targeting the *pfldh* gene (Pickard et al., 2003). Samples were then genotyped using two molecular inversion probe (MIP) panels: one targeting 1,834 variable SNP positions across the genome and another targeting known drug resistant SNP mutations in 14 genes (Aydemir et al., 2018; Verity et al., 2019). MIP capture and library preparation was done as previously described (Verity et al., 2019). Sequencing of the genome-wide (two sequencing runs) and drug resistance (one sequencing run) libraries was conducted using an Illumina NextSeq 550 instrument (150 bp paired-end reads) and an Illumina Miseq instrument (250 bp paired-end reads), respectively, at Brown University (RI, USA).

Processing of sequencing data and variant calling was done using MIPtools (v0.19.12.13; https://github.com/bailey-lab/MIPTools), a suite of computational tools designed to handle sequencing data from MIPs. Raw reads from each MIP, identifiable using unique molecular identifiers (UMIs), were used to reconstruct sequences using MIPWrangler, and variant calling was performed on these samples using freebayes (Garrison and Marth, 2012). To reduce false positives due to PCR and alignment errors, the alternative allele (SNP) must have been supported by more than one UMI within a sample, and the allele must have been represented by at least 10 UMIs across the entire population. Biallelic, variant SNP positions were retained for analysis. Low quality variants were removed from the analysis; additionally, individual variant calls within each sample were set to missing if the site was not supported by at least four UMIs. After these steps, genome positions with more than 50% missing data, and samples with subsequently more than 50% data, were removed from all downstream analyses. Variants were annotated using the 3D7 v3 reference genome.

Using the final variant set, complexity of infection (COI) for each sample was determined using the Real McCoil categorical method (Chang et al., 2017). Genetic relatedness of sample-pairs was assessed using the major allele at each position to estimate inbreeding coefficients, calculated using a maximum-likelihood approach that estimates the probability that any position is identical-by-descent between two samples (Verity et al., 2019). Networks of highly-related parasites were created using only monoclonal samples using the R igraph package (Csárdi and Nepusz, 2006). Spatial distance between sample pairs was measured both with greater circle distance and road distance. To identify genetic outliers, principal component analysis (PCA) was conducted using within-sample allele frequencies (variations using principal coordinate analysis and major alleles were also conducted with no differences in results).

## Data Availability

All sequencing data has been submitted under BioProject: pending. All code has previously been submitted to GitHub through previous publications.

## ACKNOWLEDGEMENTS

We would like to thank all of the participants who contributed to this study. We would also like to profoundly thank Dr. Steven Meshnick, who was instrumental in the inception of this work prior to his passing. We also thank Patrick Marsh for assistance in the laboratory.

