## Supplemental Figures for "Temporal and spatial analysis of *Plasmodium falciparum* genomics reveals patterns of connectivity in a low-transmission district in Southern Province, Zambia"

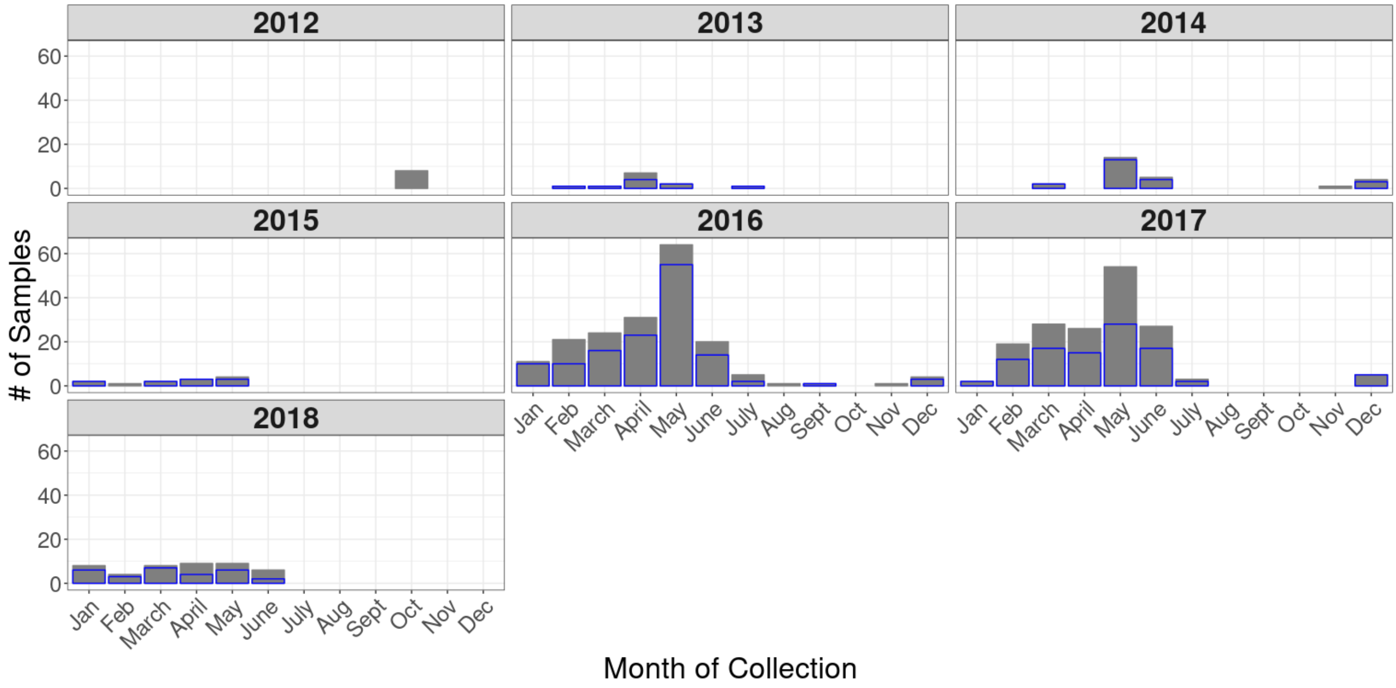


**Figure S1: Retained samples after filtering.** Total number of samples collected (in grey) are tallied by year and month of collection. Number of samples retained for analysis after filtering are shown in blue. No samples from 2012 were used for downstream analyses.

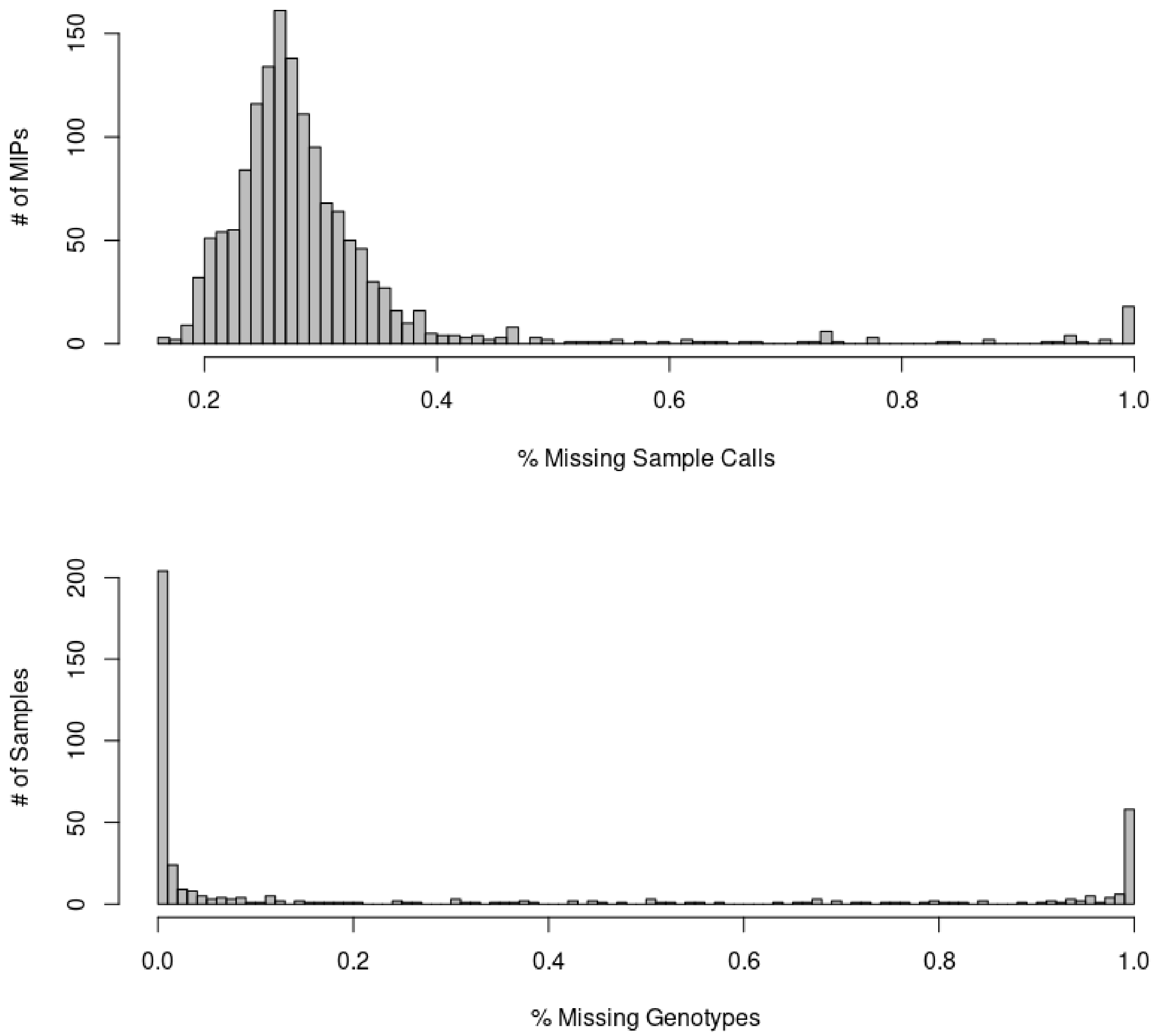


**Figure S2: Missingness by probe and sample.** Distribution of missingness across each individual targeted MIP position (top), and distribution of missingness across samples (bottom).

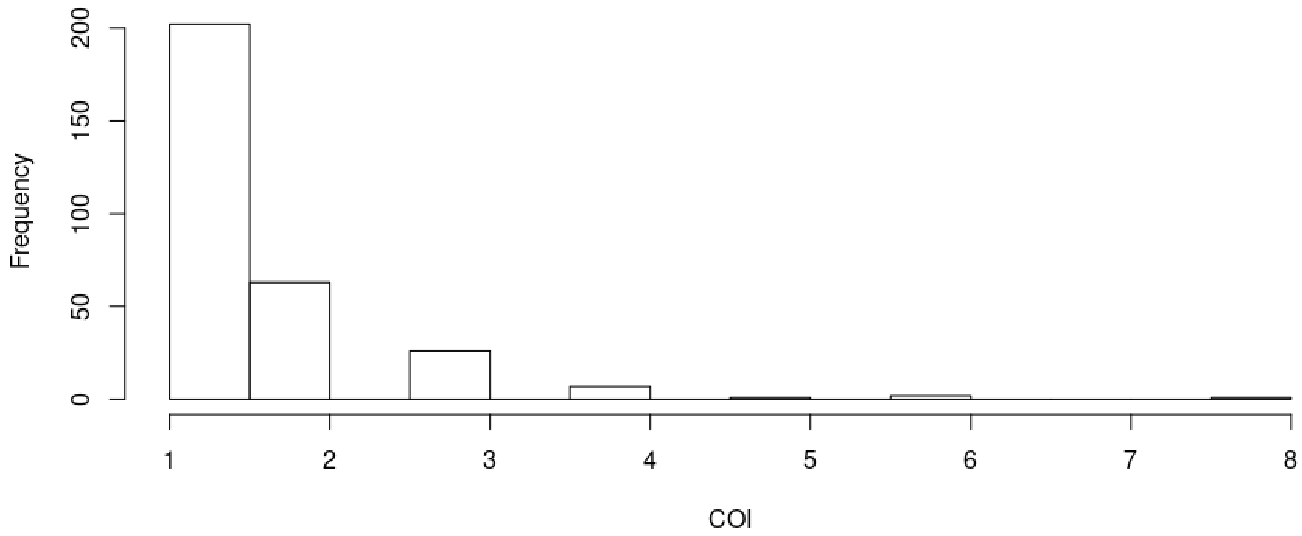


**Figure S3**: **Distribution of complexity of infection (COI) estimates from 302 genotyped infections.** The frequency of different COI determined by the RealMcCoil are shown.

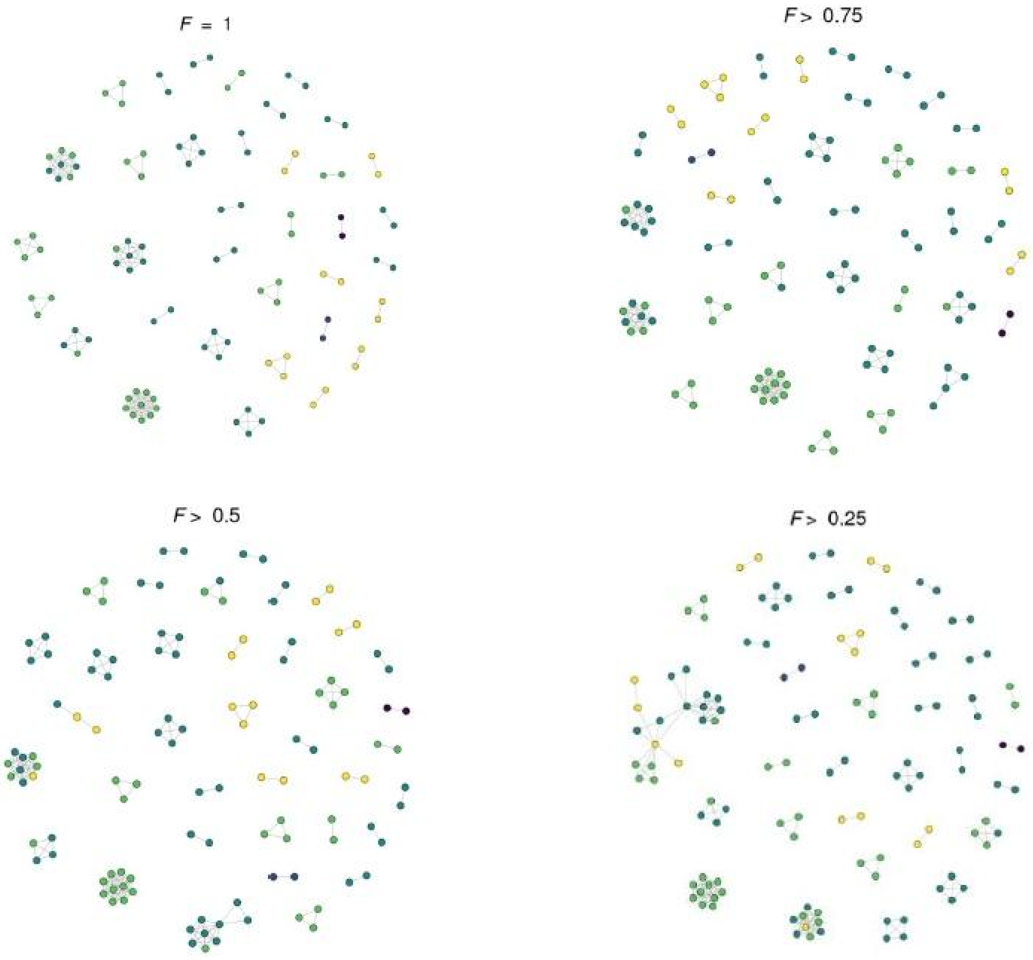


**Figure S4: Network analysis of parasite relatedness across season at varying degrees of relatedness.** Networks of monoclonal samples were generated at varying levels of genetic relatedness using the *F* statistic. The season in which a sample was collected is designated by the color of the circle. As the level of genetic relatedness decreases, increasingly complex networks begin to form and extend over multiple seasons.
